# Dexamethasone use and Mortality in Hospitalized Patients with Coronavirus Disease 2019: a Multicenter Retrospective Observational Study

**DOI:** 10.1101/2020.10.23.20218172

**Authors:** Nicolas Hoertel, Marina Sánchez, Raphaël Vernet, Nathanaël Beeker, Antoine Neuraz, Jesús Alvarado, Christel Daniel, Nicolas Paris, Alexandre Gramfort, Guillaume Lemaitre, Elisa Salamanca, Mélodie Bernaux, Ali Bellamine, Anita Burgun, Frédéric Limosin, On behalf of AP-HP / Universities / INSERM Covid-19 research collaboration and AP-HP Covid CDR Initiative

**Affiliations:** AP-HP.Centre-Université de Paris, Hôpital Corentin-Celton, Département de Psychiatrie, Issy-les-Moulineaux, France; Université de Paris, INSERM, Institut de Psychiatrie et Neurosciences de Paris, UMR_S1266, Paris, France; Université de Paris, Faculté de Santé, UFR de Médecine, Paris, France; Department of Psychobiology & Behavioural Sciences Methods, Faculty of Psychology, Universidad Complutense de Madrid, Campus de Somosaguas, Pozuelo de Alarcon, Spain; AP-HP.Centre-Université de Paris, Hôpital Européen Georges Pompidou, Medical Informatics, Biostatistics and Public Health Department, F-75015 Paris; Unité de Recherche clinique, Hopital Cochin, Assistance Publique-Hopitaux de Paris, Paris, France; INSERM, UMR_S 1138, Cordeliers Research Center, Université de Paris; Department of Medical Informatics, Necker-Enfants Malades Hospital, AP-HP.Centre-Université de Paris, 75015 Paris, France; AP-HP, DSI-WIND (Web Innovation Données), Paris, France; Sorbonne University, University Paris 13, Sorbonne Paris Cité, INSERM UMR_S 1142, F-75012 Paris, France; LIMSI, CNRS, Université Paris-Sud, Université Paris-Saclay, F-91405, Orsay, France; Université Paris-Saclay, INRIA, CEA, Palaiseau, France; Banque Nationale de Données Maladies Rares (BNDMR), Campus Picpus, Département WIND (Web Innovation Données), Paris, France; Direction de la stratégie et de la transformation, AP-HP, Paris; Unité de Recherche clinique, Hôpital Cochin, AP-HP.Centre-Université de Paris, Paris, France

**Keywords:** Covid-19, SARS-CoV-2, dexamethasone, treatment, efficacy, death, oxygen, ventilation

## Abstract

**Objective:** To examine the association between dexamethasone use and mortality among hospitalized patients for COVID-19.

**Design:** Multicenter observational retrospective cohort study.

**Setting:** Greater Paris University hospitals, France.

**Participants:** 12,217 adults hospitalized with COVID-19 between 24 January and 20 May 2020, including 171 patients (1.4%) who received dexamethasone orally or by intravenous perfusion during the visit.

**Data source:** Assistance Publique-Hôpitaux de Paris Health Data Warehouse.

**Main outcome measures:** The primary endpoint was time to death. We compared this endpoint between patients who received dexamethasone and those who did not in time-to-event analyses adjusting for sex, age, obesity, current smoking status, any medical condition associated with increased COVID-19-related mortality, and clinical and biological severity of COVID-19 at admission, while stratifying by the need of respiratory support (i.e., oxygen or intubation). The primary analysis was a multivariable Cox model and the secondary analysis used a univariate Cox regression in a matched analytic sample.

**Results:** Among patients who required respiratory support, the end-point event of death occurred in 10 patients (15.9%) who received dexamethasone and 298 patients (26.4%) who did not. In this group of patients, there was a significant association between dexamethasone use and reduced mortality in both the crude, unadjusted analysis (hazard ratio (HR), 0.40; 95% CI, 0.18 to 0.87, p=0.021) and the adjusted multivariable analysis (HR, 0.46; 95% CI, 0.22 to 0.96, p=0.039). In the sensitivity analysis, the univariate Cox regression model in the matched analytic sample yielded a same tendency, albeit non-significant (HR, 0.31; 95% CI, 0.08 to 1.14, p=0.077). Among patients without respiratory support, the end-point event of death occurred in 14 patients (13.0%) who received dexamethasone and 1,086 patients (10.0%) who did not. In this group of patients, there was no significant association between dexamethasone use and the endpoint. When examining the association between the cumulative dose of dexamethasone received during the visit and the endpoint, we found that the administration of a cumulative dose between 60 mg to 150 mg among patients who required respiratory support was significantly associated with a lower risk of death in the crude, unadjusted analysis (HR, 0.28; SE, 0.58, p=0.028), the adjusted multivariable analysis (HR, 0.24; SE, 0.65, p=0.030), and in the univariate Cox regression model in the matched analytic sample (HR, 0.32; SE, 0.58, p=0.048), whereas no significant association was observed with a different dose. Among patients without respiratory support, there was no significant association between the cumulative dose of dexamethasone and the endpoint in the crude and in the adjusted multivariable analyses.

**Conclusions:** In this observational study involving patients with Covid-19 who had been admitted to the hospital, dexamethasone use administered either orally or by intravenous injection at a cumulative dose between 60 mg and 150 mg was associated with decreased mortality among those requiring respiratory support.

## 1. Introduction

On June 16^th^, an interim analysis of the RECOVERY (Randomized Evaluation of COVid-19 thERapY) trial, a randomized clinical trial examining a range of potential treatments for COVID-19, indicated that low-dose dexamethasone could reduce mortality in patients with COVID-19 requiring oxygen or mechanical ventilation support.^1^ In that study, a total of 2,104 patients were randomized to receive dexamethasone 6 mg once per day for ten days, administered either orally or by intravenous injection, and were compared with 4,321 patients randomized to usual care alone. Dexamethasone was significantly associated with reduced 28-day mortality in ventilated patients (rate ratio (RR), 0.65; 95% confidence interval (CI), 0.48 to 0.88; p=0.0003) and in patients receiving oxygen only (RR, 0.80; 95% CI, 0.67 to 0.96; p=0.0021). No benefit was observed among patients who did not require respiratory support (RR, 1.22; 95% CI, 0.86 to 1.75; p=0.14).

These preliminary findings are of utmost importance and highlight that research into dexamethasone use in patients with COVID-19 is a priority.

In this report, we present results of a large multicenter retrospective observational study of patients admitted for Covid-19 to any of the 39 Greater Paris University hospitals. We examined whether oral or intravenous administration of dexamethasone to hospitalized adult patients with COVID-19 was associated with reduced mortality (i) among those who required respiratory support, i.e. mechanical ventilation or oxygen, and (ii) in those who did not. Following results of the RECOVERY trial interim analysis,^1^ we hypothesized that dexamethasone administration would be associated with reduced mortality in patients with COVID-19 who required respiratory support and not in those who did not.

## 2. Methods

### 2.1. Setting

We conducted this study at AP*-*HP, which comprises 39 hospitals, 23 of which are acute, 20 adult and 3 pediatric hospitals. We included all adults aged 18 years or over who have been admitted with COVID-19 to these medical centers from the beginning of the epidemic in France, i.e. January 24^th^, until May 20^th^. For all patients, COVID-19 was ascertained by a positive reverse-transcriptase–polymerase-chain-reaction (RT-PCR) test from analysis of nasopharyngeal or oropharyngeal swab specimens. This observational study using routinely collected data received approval from the Institutional Review Board of the AP-HP clinical data warehouse (decision CSE-20-20_COVID19, IRB00011591). AP-HP clinical Data Warehouse initiative ensures patients’ information and consent regarding the different approved studies through a transparency portal in accordance with European Regulation on data protection and authorization n°1980120 from National Commission for Information Technology and Civil Liberties (CNIL).

### 2.2. Data sources

We used data from the AP-HP Health Data Warehouse (‘Entrepôt de Données de Santé (EDS)’). This warehouse contains all the clinical data available on all inpatient visits for COVID-19 to any of the 39 Greater Paris University hospitals. The data obtained included patients’ demographic characteristics, vital signs, laboratory test and RT-PCR test results, medication administration data, current medication lists, current diagnoses, oxygen and ventilator use data, and death certificates.

### 2.3. Variables assessed

We obtained the following data for each patient at the time of the hospitalization: sex; age, which was categorized based on the OpenSAFELY study results^2^ (i.e. 18-50, 51-70, 71+); obesity, defined as having a body-mass index higher than 30 kg/m^2^ or an International Statistical Classification of Diseases and Related Health Problems (ICD-10) diagnosis code for obesity (E66.0, E66.1, E66.2, E66.8, E66.9); self-reported current smoking status; any medical condition associated with increased COVID-19-related mortality^2,3^ based on ICD-10 diagnosis codes, including diabetes mellitus (E11), diseases of the circulatory system (I00-I99), diseases of the respiratory system (J00-J99), neoplasms (C00-D49), and diseases of the blood and blood-forming organs and certain disorders involving the immune mechanism (D5-D8); clinical severity of COVID-19 at admission, defined as having at least one of the following criteria:^4^ (i) respiratory rate > 24 breaths/min or < 12 breaths/min, (ii) resting peripheral capillary oxygen saturation in ambient air < 90%, (iii) temperature > 40°C, or (iv) systolic blood pressure < 100 mm Hg; and biological severity of COVID-19 at admission, defined as having at least one of the following criteria:^4,5^ (i) high neutrophil-to-lymphocyte ratio or (ii) low lymphocyte-to-C-reactive protein ratio (both variables were dichotomized at the median of the values observed in the full sample), or (iii) plasma lactate levels higher than 2 mmol/L.

All medical notes and prescriptions are computerized in Greater Paris University hospitals. Medications and their mode of administration (i.e., dosage, frequency, date, condition of intake) were identified from medication administration data or scanned hand-written medical prescriptions, through two deep learning models based on BERT contextual embeddings,^6^ one for the medications and another for their mode of administration. The model was trained on the APmed corpus,^7^ a previously annotated dataset for this task. Extracted medications names were then normalized to the Anatomical Therapeutic Chemical (ATC) terminology using approximate string matching.

### 2.4. Endpoint

The endpoint was the time from study baseline to death. Patients without an end-point event had their data censored on May 20^th^, 2020.

### 2.5. Dexamethasone use

Study baseline was defined as the date of hospital admission. Dexamethasone use was defined as receiving this medication orally or by intravenous injection at any time during the follow-up period, from study baseline to the end of the hospitalization or death.

### 2.6. Dexamethasone cumulative dose

Dexamethasone cumulative dose received was calculated and considered as a categorical variable with the following categories defined *a priori*: (i) 60 mg to 150 mg based on usual prescribing practice for acute respiratory distress syndrome (ARDS) in AP-HP hospitals (corresponding to 10 mg per day for 6 days to 20 mg per day for 5 days followed by 10 mg per day for 5 days), (ii) other doses (i.e., more than 150 mg or less than 60 mg), and (iii) missing data.

### 2.7. Statistical analysis

All analyses were stratified by the need for respiratory support, i.e. oxygen or mechanical ventilation, at any time during the follow-up.

We calculated frequencies and means (± standard deviations (SD)) of each baseline characteristic described above in patients receiving and not receiving dexamethasone, and compared them using chi-square tests or Welch’s t-tests.

To examine the association of dexamethasone use and the endpoint, we performed Cox proportional-hazards regression models. To help account for the nonrandomized prescription of dexamethasone and reduce the effects of confounding, the primary analysis used a multivariable Cox regression model including as covariates sex, age, obesity, current smoking status, any medical condition associated with increased COVID-19-related mortality, and clinical and biological severity of COVID-19 at admission. Weighted Cox regression models were used when the proportional hazards assumption was not met. Kaplan-Meier curves were performed and their pointwise 95% confidence intervals were estimated using the nonparametric bootstrap method.^8^

As a sensitivity analysis, we performed a univariate Cox regression model in a matched analytic sample using a 1:10 ratio, based on the same variables used for the multivariable Cox regression analysis. To reduce the effects of confounding, optimal matching was used in order to obtain the smallest average absolute distance across all clinical characteristics between exposed patient and non-exposed matched controls.

We also examined whether the cumulative dose of dexamethasone received during the visit was associated with the endpoint.

Finally, we performed multivariable Cox regression models including interaction terms to examine whether the association between dexamethasone use and the primary endpoint significantly differed across baseline characteristics.

For all associations, we performed residual analyses to assess the fit of the data, check assumptions, including proportional hazards assumptions, and examined the potential influence of outliers. To improve the quality of result reporting, we followed the recommendations of The Strengthening the Reporting of Observational Studies in Epidemiology (STROBE) Initiative.^21^ Statistical significance was fixed *a priori* at two-sided p-value<0.05. All analyses were conducted in R software version 2.4.3 (R Project for Statistical Computing).

## 3. Results

### 3.1. Characteristics of the cohort

Of the 16,170 with a positive COVID-19 RT-PCR test consecutively admitted to the hospital from January 24^th^ to May 20^th^, a total of 3,953 patients (24.4%) were excluded because of missing data or their age (i.e. less than 18 years old of age). Of the remaining 12,217 adult inpatients, 178 patients (1.46%) received dexamethasone. Of them, 7 (3.9%) were excluded because the route of administration was either ophthalmic, through aerosol, or unknown. Among the 171 remaining patients exposed to dexamethasone, 63 (36.8%) needed respiratory support (i.e. mechanical ventilation or oxygen) and 108 (63.2%) did not (**Figure 1**). The mean cumulative dose administered was 107.8 mg (SD=63.5; median=100 mg; range: 10.0 mg to 320.0 mg) in those who required respiratory support and 101.8 mg (SD=89.8; median=90 mg; range: 10.0 mg to 600.0 mg) in those who did not. This treatment was administered orally in 97.1% of patients and by intravenous injection in 2.9% of them.

**Figure 1.**
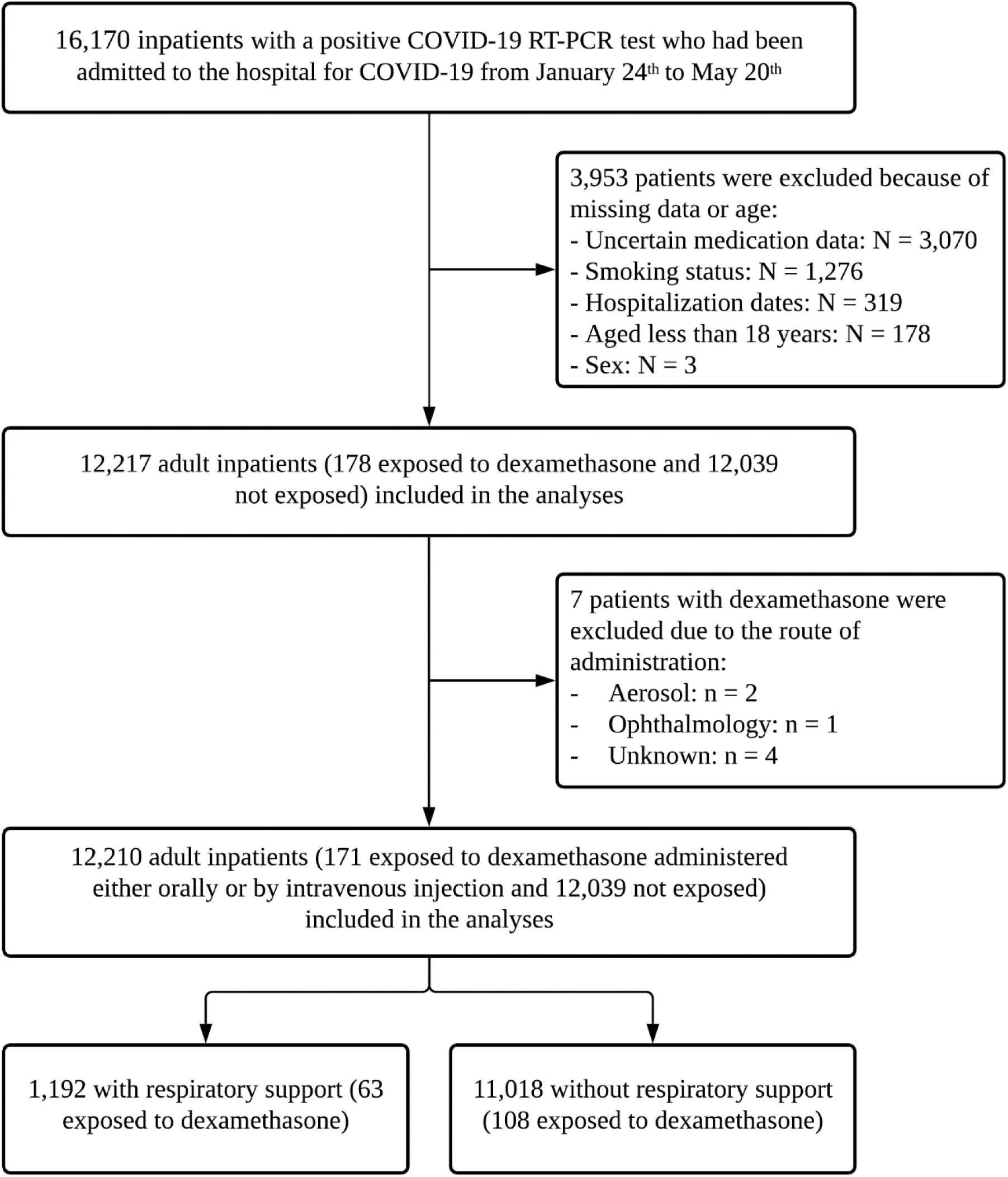
Study cohort.

COVID-19 RT-PCR test results were obtained after a mean delay of 3.7 days (SD=8.4, median=0.9 days) from the date of hospital admission in patients who required respiratory support. This delay was not significantly different between patients receiving or not receiving dexamethasone [mean delay in the exposed group=3.0 days (SD=6.2); mean delay in the non-exposed group=3.7 days (SD=8.5); Welch’s t-test=0.79, p=0.430)]. In patients who did not require respiratory support, the mean delay was 4.7 days (SD=10.2, median=1.0 day), and this delay was not significantly different between patients receiving or not receiving dexamethasone [mean delay in the exposed group=3.5 days (SD=8.9); mean delay in the non-exposed group=4.8 day (SD=10.2); Welch’s t-test=1.5, p=0.163)].

Among patients who required respiratory support, the mean follow-up was 27.9 days (SD=20.4; median=21 days; range: 1 day to 106 days) and 308 patients (25.8%) had an end-point event at the time of data cutoff on May 20^th^. Among those who did not require respiratory support, the mean follow-up was 18.5 days (SD=24.6; median=6; range: 1 day to 117 days), and 1,100 (10.0%) patients had an end-point event at the time of data cutoff.

Associations between baseline characteristics and the endpoint are given in **eTable 1**. The distribution of the patients’ characteristics by dexamethasone use is shown in **Table 1**. Dexamethasone use significantly differed in clinical and biological severity at admission among patients who required respiratory support, and in sex, age, obesity and biological severity among those who did not. In the matched analytic samples, there were no significant differences in patients’ characteristics according to dexamethasone use (**Table 1**).

**Table 1.** Characteristics of patients receiving or not receiving dexamethasone in patients with or without respiratory support (oxygen or intubation).

|  | Exposed to<br>dexamethasone | Not exposed to<br>dexamethasone | Non-exposed<br>matched group | Exposed to<br>dexamethasone<br>vs.<br>Not exposed to<br>dexamethasone<br>(crude analysis) | Exposed to<br>dexamethasone<br>vs.<br>Non-exposed matched<br>group |
| --- | --- | --- | --- | --- | --- |
|  | N (%) | N (%) |  | Chi-square test | Chi-square test<br>(p-value) |
| ***With respiratory support*** | 63 (100%) | 1,129 (100%) | 630 (100%) |  |  |
| Sex |  |  |  | <0.01 (>0.99) | 0.09 (0.770) |
| *Women* | 17 (27.0%) | 308 (27.3%) | 154 (24.4%) |  |  |
| *Men* | 46 (73.0%) | 821 (72.7%) | 476 (75.6%) |  |  |
| Age |  |  |  | 2.35 (0.309) | 0.14 (0.931) |
| *18 to 50 years* | 10 (15.9%) | 239 (21.2%) | 102 (16.2%) |  |  |
| *51 to 70 years* | 40 (63.5%) | 606 (53.7%) | 386 (61.3%) |  |  |
| *More than 70 years* | 13 (20.6%) | 284 (25.2%) | 142 (22.5%) |  |  |
| Obesity <sup>a</sup> |  |  |  | 0.25 (0.621) | <0.01 (0.946) |
| *Yes* | 16 (25.4%) | 329 (29.1%) | 168 (26.7%) |  |  |
| *No* | 47 (74.6%) | 800 (70.9%) | 462 (73.3%) |  |  |
| Smoking <sup>b</sup> |  |  |  | 0.11 (0.736) | <0.01 (0.961) |
| *Yes* | 11 (17.5%) | 170 (15.1%) | 103 (16.3%) |  |  |
| *No* | 52 (82.5%) | 959 (84.9%) | 527 (83.7%) |  |  |
| Any medical condition <sup>γ</sup> |  |  |  | 2.20 (0.138) | 1.83 (0.176) |
| *Yes* | 36 (57.1%) | 757 (67.1%) | 419 (66.5%) |  |  |
| *No* | 27 (42.9%) | 372 (32.9%) | 211 (33.5%) |  |  |
| Clinical severity of Covid-19 at admission <sup>u</sup> |  |  |  | 12.71 (0.002*) | 2.75 (0.253) |
| *Yes* | 23 (36.5%) | 529 (46.9%) | 278 (44.1%) |  |  |
| *No* | 33 (52.4%) | 354 (31.4%) | 262 (41.6%) |  |  |
| *Missing* | 7 (11.1%) | 246 (21.8%) | 90 (14.3%) |  |  |
| Biological severity of Covid-19 at admission <sup>k</sup> |  |  |  | 9.59 (0.008*) | 0.20 (0.906) |
| *Yes* | 52 (82.5%) | 716 (63.4%) | 507 (80.5%) |  |  |
| *No* | 8 (12.7%) | 279 (24.7%) | 93 (14.8%) |  |  |
| *Missing* | 3 (4.76%) | 134 (11.9%) | 30 (4.76%) |  |  |
| ***Without respiratory support*** | 108 (100%) | 10,910 (100%) | 1,080 (100%) |  |  |
| Sex |  |  |  | 21.70 (<0.001*) | 0.05 (0.820) |
| *Women* | 32 (29.6%) | 5738 (52.6%) | 337 (31.2%) |  |  |
| *Men* | 76 (70.4%) | 5172 (47.4%) | 743 (68.8%) |  |  |
| Age |  |  |  | 28.86 (<0.001*) | 0.17 (0.918) |
| *18 to 50 years* | 16 (14.8%) | 4153 (38.1%) | 147 (13.6%) |  |  |
| *51 to 70 years* | 54 (50.0%) | 3338 (30.6%) | 559 (51.8%) |  |  |
| *More than 70 years* | 38 (35.2%) | 3419 (31.3%) | 374 (34.6%) |  |  |
| Obesity <sup>a</sup> |  |  |  | 6.87 (0.009*) | <0.01 (>0.99) |
| *Yes* | 22 (20.4%) | 1279 (11.7%) | 220 (20.4%) |  |  |
| *No* | 86 (79.6%) | 9631 (88.3%) | 860 (79.6%) |  |  |
| Smoking <sup>b</sup> |  |  |  | 1.47 (0.225) | <0.01 (>0.99) |
| *Yes* | 13 (12.0%) | 908 (8.32%) | 130 (12.0%) |  |  |
| *No* | 95 (88.0%) | 10002 (91.7%) | 950 (88.0%) |  |  |
| Any medical condition <sup>y</sup> |  |  |  | 1.76 (0.184) | <0.01 (0.992) |
| *Yes* | 33 (30.6%) | 2679 (24.6%) | 336 (31.1%) |  |  |
| *No* | 75 (69.4%) | 8231 (75.4%) | 744 (68.9%) |  |  |
| Clinical severity of Covid-19 at admission <sup>u</sup> |  |  |  | 5.49 (0.064) | <0.01 (0.998) |
| *Yes* | 17 (15.7%) | 2022 (18.5%) | 170 (15.7%) |  |  |
| *No* | 38 (35.2%) | 2763 (25.3%) | 377 (34.9%) |  |  |
| *Missing* | 53 (49.1%) | 6125 (56.1%) | 533 (49.4%) |  |  |
| Biological severity of Covid-19 at admission <sup>k</sup> |  |  |  | 61.83 (<0.001*) | 0.01 (0.993) |
| *Yes* | 69 (63.9%) | 3254 (29.8%) | 696 (64.4%) |  |  |
| *No* | 22 (20.4%) | 2903 (26.6%) | 216 (20.0%) |  |  |
| *Missing* | 17 (15.7%) | 4753 (43.6%) | 168 (15.6%) |  |  |

<sup>a</sup> Defined as having a body-mass index higher than 30 kg/m<sup>2</sup> or an International Statistical Classification of Diseases and Related Health Problems (ICD-10) diagnosis code for obesity (E66.0, E66.1, E66.2, E66.8, E66.9).
<sup>β</sup> Current smoking status was self-reported.
<sup>γ</sup> Assessed using ICD-10 diagnosis codes for diabetes mellitus (E11), diseases of the circulatory system (I00-I99), diseases of the respiratory system (J00-J99), neoplasms (C00-D49), and diseases of the blood and blood-forming organs and certain disorders involving the immune mechanism (D5-D8)
<sup>u</sup> Defined as having at least one of the following criteria: respiratory rate > 24 breaths/min or < 12 breaths/min, resting peripheral capillary oxygen saturation in ambient air < 90%, temperature > 40°C, or systolic blood pressure < 100 mm Hg.
<sup>k</sup> Defined as having at least one of the following criteria: high neutrophil-to-lymphocyte ratio, low lymphocyte-to-C-reactive protein (both variables were dichotomized at the median of the values observed in the full sample), and plasma lactate levels higher than 2 mmol/L.
\* p-value is significant (p<0.05).

### 3.2. Study endpoint

Among patients who required respiratory support, the end-point event of death occurred in 10 patients (15.9%) who received dexamethasone and 298 patients (26.4%) who did not (**Table 2**). In this group, we found a significant association between dexamethasone use and reduced risk of the endpoint of death in both the crude, unadjusted analysis (hazard ratio (HR), 0.40; 95% CI, 0.18 to 0.87, p=0.021) and the adjusted multivariable analysis (HR, 0.46; 95% CI, 0.22 to 0.96, p=0.039) (**Figure 2**; **Table 2**). In the sensitivity analysis, the univariate Cox regression model in the matched analytic sample yielded a same tendency, albeit non-significant (HR, 0.31; 95% CI, 0.08 to 1.14, p=0.077) (**Figure 3**; **Table 2**).

**Table 2.**
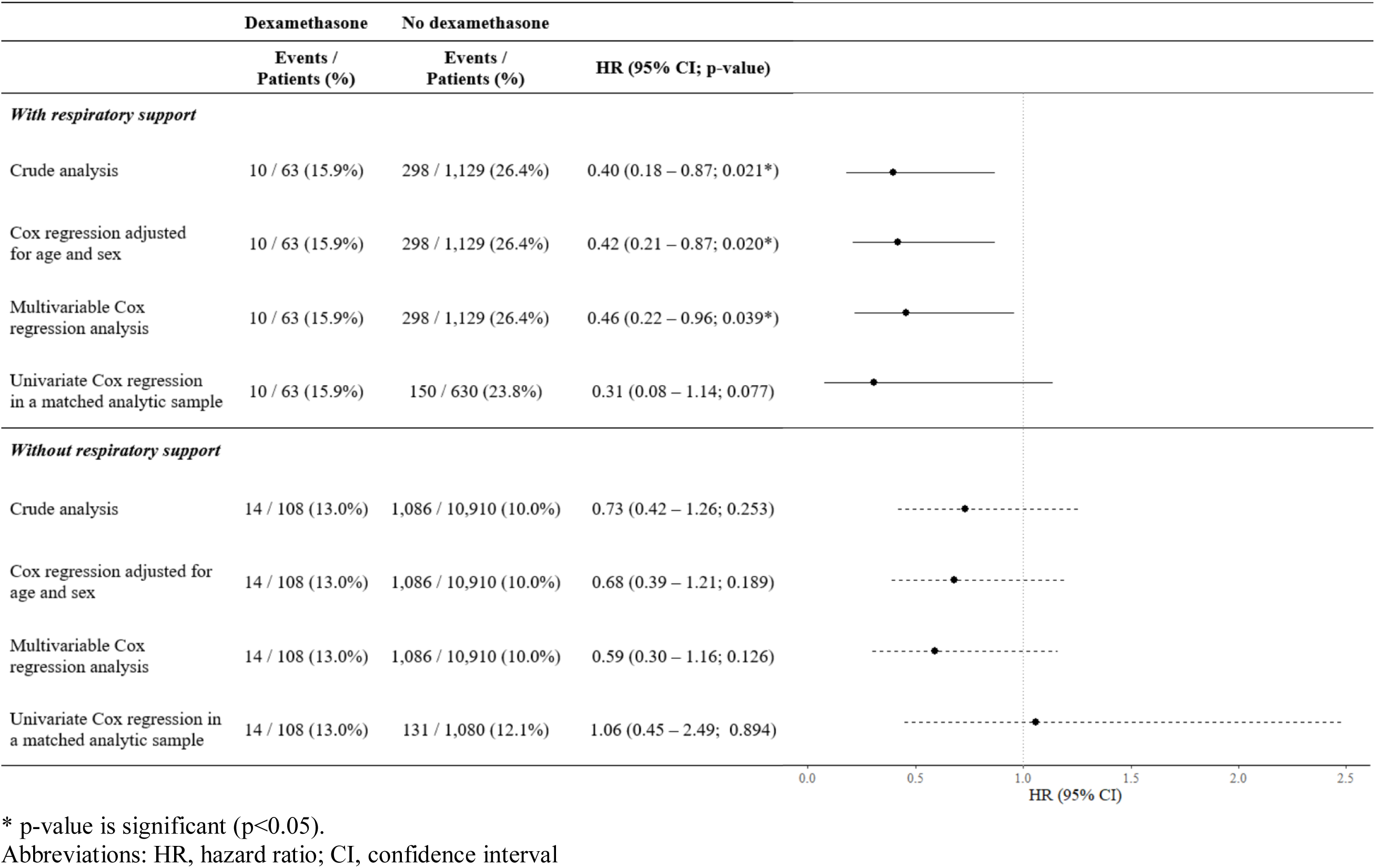
Association between dexamethasone use and the endpoint of death in the full sample and in the matched analytic sample.

|  | Dexamethasone | No dexamethasone |  |
| --- | --- | --- | --- |
|  | Events /<br>Patients (%) | Events /<br>Patients (%) | HR (95% CI; p-value) |
| *With respiratory support* |  |  |  |
| Crude analysis | 10 / 63 (15.9%) | 298 / 1,129 (26.4%) | 0.40 (0.18 – 0.87; 0.021*) |
| Cox regression adjusted for age and sex | 10 / 63 (15.9%) | 298 / 1,129 (26.4%) | 0.42 (0.21 – 0.87; 0.020*) |
| Multivariable Cox regression analysis | 10 / 63 (15.9%) | 298 / 1,129 (26.4%) | 0.46 (0.22 – 0.96; 0.039*) |
| Univariate Cox regression in a matched analytic sample | 10 / 63 (15.9%) | 150 / 630 (23.8%) | 0.31 (0.08 – 1.14; 0.077) |
| *Without respiratory support* |  |  |  |
| Crude analysis | 14 / 108 (13.0%) | 1,086 / 10,910 (10.0%) | 0.73 (0.42 – 1.26; 0.253) |
| Cox regression adjusted for age and sex | 14 / 108 (13.0%) | 1,086 / 10,910 (10.0%) | 0.68 (0.39 – 1.21; 0.189) |
| Multivariable Cox regression analysis | 14 / 108 (13.0%) | 1,086 / 10,910 (10.0%) | 0.59 (0.30 – 1.16; 0.126) |
| Univariate Cox regression in a matched analytic sample | 14 / 108 (13.0%) | 131 / 1,080 (12.1%) | 1.06 (0.45 – 2.49; 0.894) |

\* p-value is significant (p<0.05).
Abbreviations: HR, hazard ratio; CI, confidence interval

**Figure 2.**
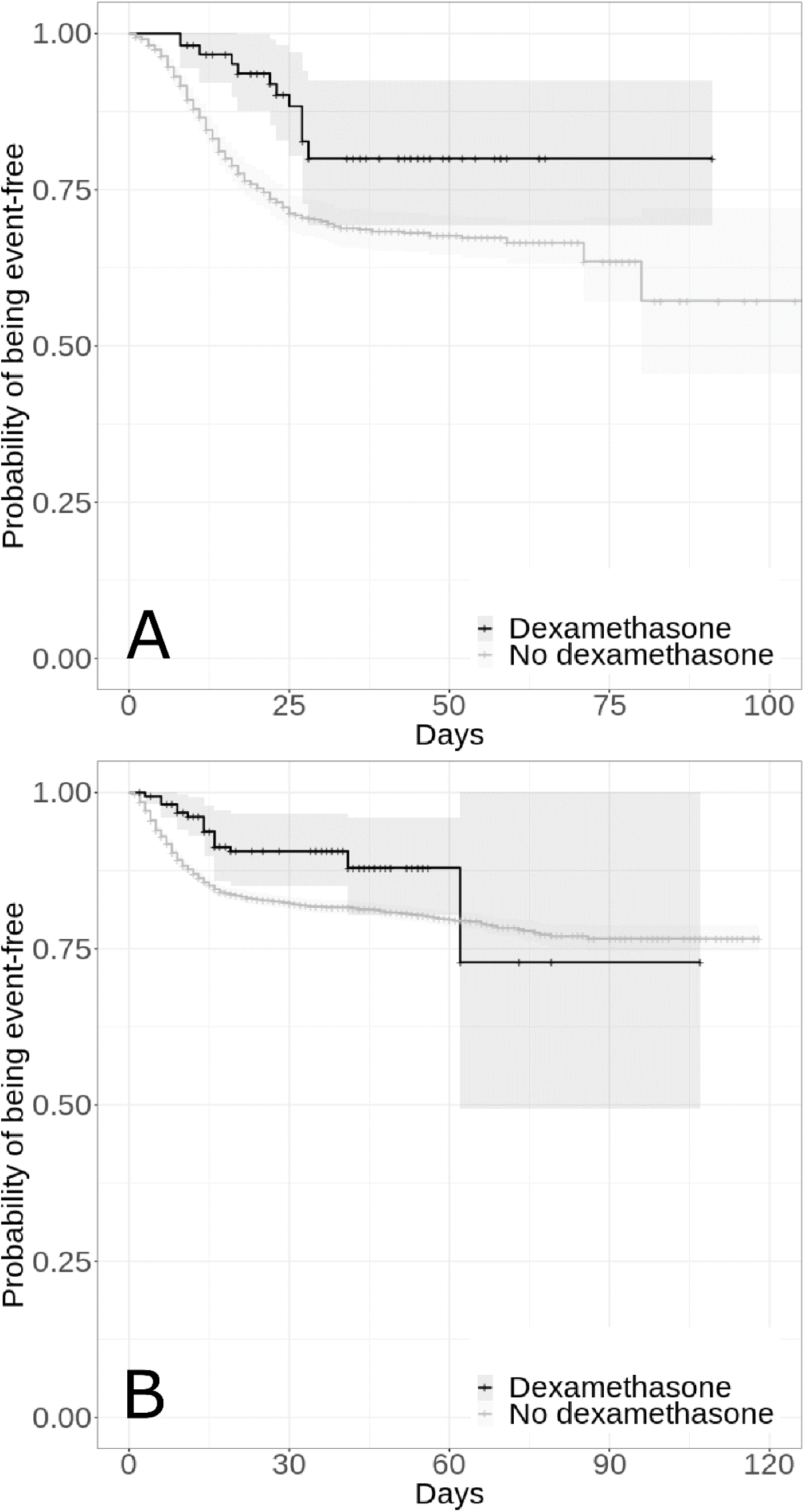
Kaplan-Meier curves for death in the full samples of hospitalized patients with Covid-19 who required respiratory support (i.e., mechanical ventilation or oxygen) (N=1,192) (A), and of those who did not (N=11,018) (B), according to dexamethasone use. The shaded areas represent pointwise 95% confidence intervals.

**Figure 3.**
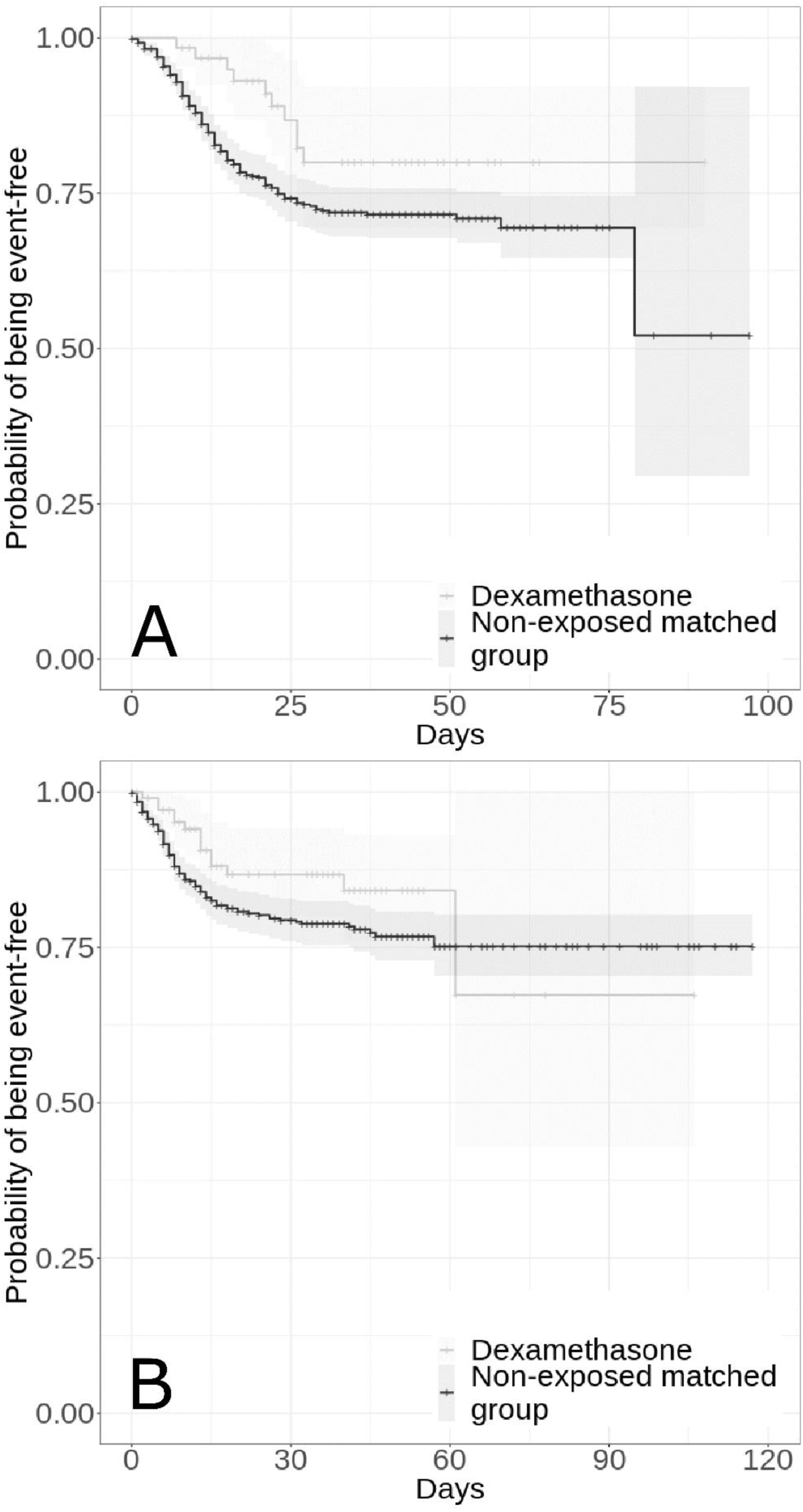
Kaplan-Meier curves for death in the matched analytic samples of hospitalized patients with Covid-19 who required respiratory support (i.e., mechanical ventilation or oxygen) (N=693) (A) and of those who did not (N=1,188) (B) according to dexamethasone use. The shaded areas represent pointwise 95% confidence intervals. For each exposed case, ten non-exposed controls were selected.

Among patients without respiratory support, the end-point event of death occurred in 14 patients (13.0%) who received dexamethasone and 1,086 patients (10.0%) who did not (**Table 2**). In this group of patients, there was no significant association between dexamethasone use and the endpoint, neither in the crude, unadjusted analysis (HR, 0.73; 95% CI, 0.42 to 1.26, p=0.253) or in the adjusted multivariable analysis (HR, 0.59; 95% CI, 0.30 to 1.16, p=0.126) (**Figure 2**; **Table 2**). In the sensitivity analysis, the univariate Cox regression model in the matched analytic sample yielded a similar result (HR, 1.06; 95% CI, 0.45 to 2.49, p=0.894) (**Figure 3**; **Table 2**).

A post-hoc analysis indicated that in the full sample of patients with respiratory support, we had 80% power to detect hazard ratios for dexamethasone treatment of at least 2.33/0.15 and of at least 2.37/0.14 in the matched analytic sample. In those without respiratory support, we had 80% power to detect hazard ratios for dexamethasone treatment of at least 1.99/0.30 in the full sample and 2.05/0.29 in the matched analytic sample.

When examining the association between the cumulative dose of dexamethasone received during the visit and the endpoint, we found that the administration of a cumulative dose between 60 mg to 150 mg among patients who required respiratory support was significantly associated with a lower risk of death in the crude, unadjusted analysis (HR, 0.28; SE, 0.58, p=0.028), the adjusted multivariable analysis (HR, 0.24; SE, 0.65, p=0.030), and in the univariate Cox regression model in the matched analytic sample (HR, 0.32; SE, 0.58, p=0.048), whereas no significant association was observed with a different dose (**eTable 2, eFigure 1**). Among patients without respiratory support, there was no significant association between the cumulative dose of dexamethasone and the endpoint in the crude, unadjusted analysis (HR, 0.37; SE, 0.58, p=0.089) and the adjusted multivariable analysis (HR, 0.47; SE, 0.57, p=0.178). However, the administration of a cumulative dose between 60 mg to 150 mg was significantly associated with the endpoint in the univariate Cox regression model in the matched analytic sample (HR, 0.32; SE, 0.58, p=0.049) (**eTable 2, eFigure 1**).

Finally, the association between dexamethasone use and the outcome did not significantly differ across subgroups defined by baseline characteristics in both groups with and without respiratory support, except for patients with obesity who did not require respiratory support, for whose dexamethasone use was significantly associated with a higher risk of death compared to obese patients without dexamethasone (HR, 3.90; 95% CI, 1.13 to 13.44, p=0.031) (**eTable 3**). However, none of the 16 patients with obesity who received a cumulative dose of 60 to 150 mg of dexamethasone died.

## 4. Discussion

In this multicenter retrospective observational study involving a large sample of patients admitted to the hospital with COVID-19, we found that dexamethasone use, administered either orally or by intravenous injection at a cumulative dose between 60 mg and 150 mg, was significantly and substantially associated with reduced mortality among patients with COVID-19 requiring oxygen or mechanical ventilation support. This association did not significantly differ according to baseline clinical characteristics. No significant association between dexamethasone use and mortality was observed among patients with COVID-19 without respiratory support, except in the univariate Cox regression model in the matched analytic sample where the administration of a cumulative dose between 60 mg to 150 mg was significantly associated with reduced mortality. Although these findings should be interpreted with caution due to the observational design, they are in line with the results of the RECOVERY trial interim analysis,^1^ which indicated that dexamethasone 6 mg once per day for ten days, administered either orally or by intravenous injection, was significantly associated with reduced 28-day mortality in ventilated patients and in patients receiving oxygen only, and extend them by suggesting that cumulative dose between 60 mg and 150 mg might be efficient in reducing mortality among patients with COVID-19 requiring respiratory support.

Our study has several limitations. First, some amount of unmeasured confounding may remain. However, our analyses adjusted for numerous potential confounders, including sex, age, obesity, current smoking status, any medical condition associated with increased COVID-19-related mortality, and clinical and biological severity of COVID-19 at admission. Second, there are missing data for some variables and potential for inaccuracies in the electronic health records, such as the possible lack of documentation of illnesses or medications, or the misidentification of treatment mode of administration (e.g., dose, frequency), especially for hand-written medical prescriptions. Finally, despite the multicenter design, our results may not be generalizable to other settings or regions.

In this observational study involving patients with Covid-19 who had been admitted to the hospital, dexamethasone use administered either orally or by intravenous injection at a cumulative dose between 60 mg and 150 mg was associated with decreased mortality among patients requiring respiratory support.

## Supporting information

eFigure 1

eTable 1

eTable 2

eTable 3

## Data Availability

Data from the AP-HP Health Data Warehouse can be obtained at https://eds.aphp.fr//.

## Acknowledgments

The authors thank the EDS APHP Covid consortium integrating the APHP Health Data Warehouse team as well as all the APHP staff and volunteers who contributed to the implementation of the EDS-Covid database and operating solutions for this database.

Collaborators of the EDS APHP Covid consortium are: Pierre-Yves ANCEL, Alain BAUCHET, Nathanaël BEEKER, Vincent BENOIT, Mélodie BERNAUX, Ali BELLAMINE, Romain BEY, Aurélie BOURMAUD, Stéphane BREANT, Anita BURGUN, Fabrice CARRAT, Charlotte CAUCHETEUX, Julien CHAMP, Sylvie CORMONT, Christel DANIEL, Julien DUBIEL, Catherine DUCLOAS, Loic ESTEVE, Marie FRANK, Nicolas GARCELON, Alexandre GRAMFORT, Nicolas GRIFFON, Olivier GRISEL, Martin GUILBAUD, Claire HASSEN-KHODJA, François HEMERY, Martin HILKA, Anne-Sophie JANNOT, Jerome LAMBERT, Richard LAYESE, Judith LEBLANC, Léo LEBOUTER, Guillaume LEMAITRE, Damien LEPROVOST, Ivan LERNER, Kankoe LEVI SALLAH, Aurélien MAIRE, Marie-France MAMZER, Patricia MARTEL, Arthur MENSCH, Thomas MOREAU, Antoine NEURAZ, Nina ORLOVA, Nicolas PARIS, Bastien RANCE, Hélène RAVERA, Antoine ROZES, Elisa SALAMANCA, Arnaud SANDRIN, Patricia SERRE, Xavier TANNIER, Jean-Marc TRELUYER, Damien VAN GYSEL, Gaël VAROQUAUX, Jill Jen VIE, Maxime WACK, Perceval WAJSBURT, Demian WASSERMANN, Eric ZAPLETAL.

## Notes

### Competing Interest Statement

Dr Hoertel has received personal fees and non-financial support from Lundbeck, outside the submitted work. Dr Limosin has received speaker and consulting fees from Janssen-Cilag outside the submitted work. Other authors declare no competing interests.

### Funding Statement

This work did not receive any external funding.

### Author Declarations

This observational study using routinely collected data received approval from the Institutional Review Board of the AP-HP clinical data warehouse (decision CSE-20-20_COVID19, IRB00011591). AP-HP clinical Data Warehouse initiative ensures patient information and consent regarding the different approved studies and data opt-out service through a transparency portal in accordance with European Regulation on data protection and authorization number 1980120 from National Commission for Information Technology and Civil Liberties (CNIL). All procedures related to this work adhered to the ethical standards of the relevant national and institutional committees on human experimentation and with the Helsinki Declaration of 1975, as revised in 2008.

