## Supplementary material for "Dexamethasone use and Mortality in Hospitalized Patients with Coronavirus Disease 2019: a Multicenter Retrospective Observational Study": eFigure 1

**eFigure 1. Freedom from the composite endpoint of intubation or death in patients with oxygen or intubation (N=693) (A) and in patients without oxygen or intubation (N=1,043) (B) of hospitalized patients with Covid-19 according to dexamethasone dosage, in the matched sample.**

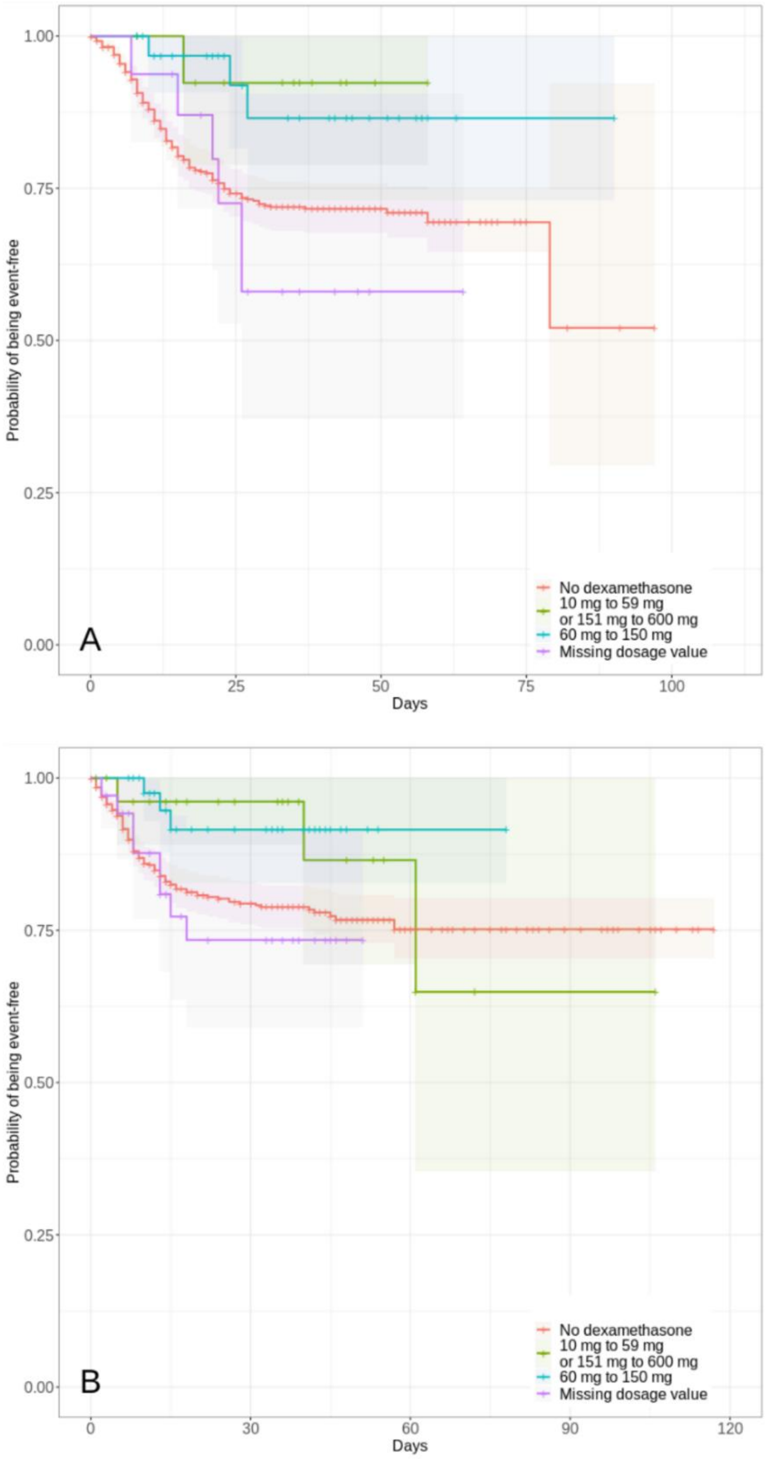


*Note*: The shaded areas represent pointwise 95% confidence intervals.
