## Supplementary material for "Dexamethasone use and Mortality in Hospitalized Patients with Coronavirus Disease 2019: a Multicenter Retrospective Observational Study": eTable 1

**eTable 1. Associations of baseline clinical characteristics with the endpoint of death in patients with and without respiratory support (oxygen or intubation).**

| | Full population | With the end-point event | Without the end-point event | Crude analysis | Mutivariable analysis | |
|---|---|---|---|---|---|---|
| | N (%) | N (%) | N (%) | HR (SE) / p-value | HR (SE) / p-value | Collinearity diagnosis (VIF) |
| ***With respiratory support*** | 1,192 (100%) | 308 (25.8%) | 884 (74.2%) | | | |
| Sex | | | | | | 1.02 |
| *Women* | 325 (27.3%) | 78 (25.3%) | 247 (27.9%) | Ref. | Ref. | |
| *Men* | 867 (72.7%) | 230 (74.7%) | 637 (72.1%) | 1.08 (0.13) / 0.563 | 1.23 (0.18) / 0.254 | |
| Age | | | | | | 1.01 |
| *18 to 50 years* | 249 (20.9%) | 32 (10.4%) | 217 (24.5%) | Ref. | Ref. | |
| *51 to 70 years* | 646 (54.2%) | 149 (48.4%) | 497 (56.2%) | 1.73 (0.19) / 0.005* | 1.03 (0.36) / 0.934 | |
| *More than 70 years* | 297 (24.9%) | 127 (41.2%) | 170 (19.2%) | 3.79 (0.20) / <0.001* | 2.92 (0.38) / 0.005* | |
| Obesity $^{\alpha}$ | | | | | | 1.02 |
| *Yes* | 345 (28.9%) | 92 (29.9%) | 253 (28.6%) | 1.04 (0.12) / 0.735 | 0.83 (0.21) / 0.379 | |
| *No* | 847 (71.1%) | 216 (70.1%) | 631 (71.4%) | Ref. | Ref. | |
| Smoking $^{\beta}$ | | | | | | 1.01 |
| *Yes* | 181 (15.2%) | 38 (12.3%) | 143 (16.2%) | 0.75 (0.17) / 0.095 | 0.60 (0.35) / 0.142 | |
| *No* | 1011 (84.8%) | 270 (87.7%) | 741 (83.8%) | Ref. | Ref. | |
| Any medical condition $^{\gamma}$ | | | | | | 1.03 |
| *Yes* | 793 (66.5%) | 257 (83.4%) | 536 (60.6%) | 3.47 (0.15) / <0.001* | 4.35 (0.20) / <0.001* | |
| *No* | 399 (33.5%) | 51 (16.6%) | 348 (39.4%) | Ref. | Ref. | |
| Clinical severity of Covid-19 at admission $^{\mu}$ | | | | | | 1.02 |

| | | | | | | |
|---|---|---|---|---|---|---|
| *Yes* | 552 (46.3%) | 147 (47.7%) | 405 (45.8%) | 1.21 (0.13) / 0.157 | 1.53 (0.17) / 0.011* | |
| *No* | 387 (32.5%) | 89 (28.9%) | 298 (33.7%) | Ref. | Ref. | |
| *Missing* | 253 (21.2%) | 72 (23.4%) | 181 (20.5%) | 1.38 (0.16) / 0.041* | 1.92 (0.23) / 0.004* | |
| Biological severity of Covid-19 at admission [κ] | | | | | | 1.01 |
| *Yes* | 768 (64.4%) | 193 (62.7%) | 575 (65.0%) | 0.89 (0.13) / 0.390 | 1.06 (0.20) / 0.778 | |
| *No* | 287 (24.1%) | 79 (25.6%) | 208 (23.5%) | Ref. | Ref. | |
| *Missing* | 137 (11.5%) | 36 (11.7%) | 101 (11.4%) | 1.00 (0.20) / 0.998 | 1.20 (0.37) / 0.654 | |
| ***Without respiratory support*** | 11,018 (100%) | 1,100 (10.0%) | 9,918 (90.0%) | | | |
| Sex | | | | | | 1.03 |
| *Women* | 5770 (52.4%) | 450 (40.9%) | 5320 (53.6%) | Ref. | Ref. | |
| *Men* | 5248 (47.6%) | 650 (59.1%) | 4598 (46.4%) | 1.50 (0.06) / <0.001* | 1.14 (0.09) / 0.134 | |
| Age | | | | | | 1.08 |
| *18 to 50 years* | 4169 (37.8%) | 19 (1.73%) | 4150 (41.8%) | Ref. | Ref. | |
| *51 to 70 years* | 3392 (30.8%) | 159 (14.5%) | 3233 (32.6%) | 8.18 (0.24) / <0.001* | 4.46 (0.35) / <0.001* | |
| *More than 70 years* | 3457 (31.4%) | 922 (83.8%) | 2535 (25.6%) | 35.71 (0.23) / <0.001* | 21.42 (0.34) / <0.001* | |
| Obesity [α] | | | | | | 1.02 |
| *Yes* | 1301 (11.8%) | 200 (18.2%) | 1101 (11.1%) | 1.36 (0.08) / <0.001* | 1.09 (0.10) / 0.403 | |
| *No* | 9717 (88.2%) | 900 (81.8%) | 8817 (88.9%) | Ref. | Ref. | |
| Smoking [β] | | | | | | 1.03 |
| *Yes* | 921 (8.36%) | 166 (15.1%) | 755 (7.61%) | 1.48 (0.08) / <0.001* | 0.93 (0.10) / 0.485 | |
| *No* | 10097 (91.6%) | 934 (84.9%) | 9163 (92.4%) | Ref. | Ref. | |
| Any medical condition [γ] | | | | | | 1.14 |
| *Yes* | 2712 (24.6%) | 637 (57.9%) | 2075 (20.9%) | 4.66 (0.06) / <0.001* | 3.19 (0.09) / <0.001* | |
| *No* | 8306 (75.4%) | 463 (42.1%) | 7843 (79.1%) | Ref. | Ref. | |

| | | | | | | |
|---|---|---|---|---|---|---|
| Clinical severity of Covid-19 at admission [μ] | | | | | | 1.13 |
| *Yes* | 2039 (18.5%) | 480 (43.6%) | 1559 (15.7%) | 2.53 (0.08) / <0.001* | 2.07 (0.11) / <0.001* | |
| *No* | 2801 (25.4%) | 260 (23.6%) | 2541 (25.6%) | Ref. | Ref. | |
| *Missing* | 6178 (56.1%) | 360 (32.7%) | 5818 (58.7%) | 0.75 (0.08) / <0.001* | 1.75 (0.13) / <0.001* | |
| Biological severity of Covid-19 at admission [κ] | | | | | | 1.14 |
| *Yes* | 3323 (30.2%) | 670 (60.9%) | 2653 (26.7%) | 2.54 (0.08) / <0.001* | 1.72 (0.10) / <0.001* | |
| *No* | 2925 (26.5%) | 243 (22.1%) | 2682 (27.0%) | Ref. | Ref. | |
| *Missing* | 4770 (43.3%) | 187 (17.0%) | 4583 (46.2%) | 0.61 (0.10) / <0.001* | 1.15 (0.14) / 0.321 | |

[α] Defined as having a body-mass index higher than 30 $kg/m^2$ or an International Statistical Classification of Diseases and Related Health Problems (ICD-10) diagnosis code for obesity (E66.0, E66.1, E66.2, E66.8, E66.9).
[β] Current smoking status was self-reported.
[γ] Assessed using ICD-10 diagnosis codes for diabetes mellitus (E11), diseases of the circulatory system (I00-I99), diseases of the respiratory system (J00-J99), neoplasms (C00-D49), and diseases of the blood and blood-forming organs and certain disorders involving the immune mechanism (D5-D8)
[μ] Defined as having at least one of the following criteria: respiratory rate > 24 breaths/min or < 12 breaths/min, resting peripheral capillary oxygen saturation in ambient air < 90%, temperature > 40°C, or systolic blood pressure < 100 mm Hg.
[κ] Defined as having at least one of the following criteria: high neutrophil-to-lymphocyte ratio, low *lymphocyte*-to-*C-reactive protein* (both variables were dichotomized at the median of the values observed in the full sample), and plasma *lactate* levels *higher than 2 mmol/L*.
* p-value is significant (p<0.05).
Abbreviations: HR, hazard ratio; SE, standard error; VIF, variance inflation factor.
