## Supplementary material for "Dexamethasone use and Mortality in Hospitalized Patients with Coronavirus Disease 2019: a Multicenter Retrospective Observational Study": eTable 2

**eTable 2. Associations between dexamethasone dosage and the endpoint of death in the full sample and in the matched analytic sample.**

| With oxygen or intubation | | | | | Without oxygen or intubation | | | | |
|---|---|---|---|---|---|---|---|---|---|
| **Dexamethasone total dosage** | **Crude analysis** | **Cox regression adjusted for age and sex** | **Multivariable (Weighted) Cox regression analysis** | **Univariate Cox regression in the matched analytic samples** | **Dexamethasone total dosage** | **Crude analysis** | **Cox regression adjusted for age and sex** | **Multivariable (Weighted) Cox regression analysis** | **Univariate Cox regression in the matched analytic samples** |
| | **HR (SE) / p-value** | **HR (SE) / p-value** | **HR (SE) / p-value** | **HR (SE) / p-value** | | **HR (SE) / p-value** | **HR (SE) / p-value** | **HR (SE) / p-value** | **HR (SE) / p-value** |
| No dexamethasone (n=1,129) | Ref. | Ref. | Ref. | Ref. | No dexamethasone (n=10,910) | Ref. | Ref. | Ref. | Ref. |
| Other dosage (n=14) | 0.21 (1.00) / 0.117 | 0.21 (1.00) / 0.115 | 0.24 (1.03) / 0.169 | 0.24 (1.00) / 0.150 | Other dosage (n=18) | 0.60 (0.58) / 0.376 | 0.57 (0.58) / 0.332 | 0.23 (0.90) / 0.101 | 0.50 (0.58) / 0.241 |
| 60 mg to 150 mg (n =33) | 0.28 (0.58) / 0.028* | 0.28 (0.58) / 0.027* | 0.24 (0.65) / 0.030* | 0.32 (0.58) / 0.048* | 60 mg to 150 mg (n=45) | 0.37 (0.58) / 0.089 | 0.41 (0.58) / 0.122 | 0.47 (0.57) / 0.178 | 0.32 (0.58) / 0.049* |
| Missing (n=16) | 1.16 (0.41) / 0.723 | 1.22 (0.41) / 0.636 | 1.10 (0.51) / 0.856 | 1.32 (0.42) / 0.507 | Missing (n=35) | 1.40 (0.35) / 0.355 | 1.19 (0.36) / 0.618 | 1.32 (0.31) / 0.361 | 1.19 (0.36) / 0.634 |

* p-value is significant (p<0.05).
Abbreviations: HR, hazard ratio; SE, standard error.
