## Supplementary material for "Dexamethasone use and Mortality in Hospitalized Patients with Coronavirus Disease 2019: a Multicenter Retrospective Observational Study": eTable 3

**eTable 3. Interaction of baseline characteristics with dexamethasone use on the endpoint of death among adult inpatients with COVID-19.**

| | With respiratory support | Without respiratory support |
|---|---|---|
| | **Multivariable Cox regression analysis** | **Multivariable Cox regression analysis** |
| | **HR (SE) / p-value** | **HR (SE) / p-value** |
| *Characteristics* | | |
| Sex | | |
| *Women* | Ref. | Ref. |
| *Men* | 1.86 (0.22 – 15.99; 0.573) | 0.91 (0.18 – 4.57; 0.907) |
| Age | | |
| *18 to 50 years* | Ref. | Ref. |
| *51 to 70 years* | 0.64 (0.11 – 3.66; 0.619) | NA |
| *More than 70 years* | 0.24 (0.03 – 1.84; 0.172) | NA |
| Obesity [α] | | |
| *Yes* | 2.54 (0.59 – 11.02; 0.213) | 3.90 (1.13 – 13.44; 0.031*) |
| *No* | Ref. | Ref. |
| Smoking | | |
| *Yes* | 1.28 (0.12 – 13.51; 0.839) | 0.22 (0.02 – 2.64; 0.233) |
| *No* | Ref. | Ref. |
| Any medical condition [β] | | |
| *Yes* | 0.35 (0.08 – 1.62; 0.181) | 1.11 (0.28 – 4.34; 0.880) |
| *No* | Ref. | Ref. |
| Clinical severity of Covid-19 at admission [μ] | | |
| *Yes* | 1.35 (0.30 – 6.05; 0.694) | 0.30 (0.04 – 2.55; 0.788) |
| *No* | Ref. | Ref. |
| *Missing* | NA | 0.46 (0.12 – 1.78; 0.263) |
| Biological severity of Covid-19 at admission [κ] | | |
| *Yes* | 0.69 (0.10 – 4.70; 0.701) | 0.31 (0.09 – 1.01; 0.052) |
| *No* | Ref. | Ref. |
| *Missing* | 3.24 (0.30 – 35.00; 0.332) | 0.35 (0.04 – 3.35; 0.362) |

[α] Defined as having a body-mass index higher than 30 kg/m$^2$ or an International Statistical Classification of Diseases and Related Health Problems (ICD-10) diagnosis code for obesity (E66.0, E66.1, E66.2, E66.8, E66.9).
[β] Assessed using ICD-10 codes for diabetes mellitus, diseases of the circulatory system, diseases of the respiratory system, neoplasms, and diseases of the blood and blood-forming organs and certain disorders involving the immune mechanism based on ICD-10 classification.
[μ] Defined as having at least one of the following criteria: respiratory rate > 24 breaths/min, oxygen saturation < 92%, temperature ≥ 40°C, and systolic blood pressure < 100 mm Hg.
[κ] Defined as having at least one of the following criteria: neutrophil-to-lymphocyte ratio higher than the median in the full sample, *lymphocyte*-to-*C-reactive protein ratio* lower than the median in the full sample, and plasma *lactate* level *higher than 2 mmol/L.*
* p-value is significant (p<0.05).
